# The seasonality of cholera in sub-Saharan Africa

**DOI:** 10.1101/2021.11.23.21265642

**Authors:** Javier Perez-Saez, Justin Lessler, Elizabeth C. Lee, Francisco J. Luquero, Espoir Bwenge Malembaka, Flavio Finger, José Paulo Langa, Sebastian Yennan, Ben Zaitchik, Andrew S Azman

## Abstract

**Background:** Cholera remains a major threat in Sub-Saharan Africa (SSA) where some of the highest case fatality risks are reported. Knowing in what months and where cholera tends to occur across the continent can aid in improving efforts to eliminate cholera as a public health concern; though largely due to lack of unified large-scale datasets, no continent-wide estimates exist. In this study we aim to estimate cholera seasonality across SSA.

**Methods:** We leverage the Global Task Force on Cholera Control (GTFCC) global cholera database with statistical models to synthesize data across spatial and temporal scale in order to infer the seasonality of excess suspected cholera occurrence in SSA. We developed a Bayesian statistical model to infer the monthly risk of excess cholera at the first and/or second administrative levels. Seasonality patterns were then grouped into spatial clusters. Finally, we studied the association between seasonality estimates and hydro-climatic variables.

**Findings:** The majority of studied countries (24/34) have seasonal patterns in excess cholera, corresponding to approximately 85% of the SSA population. Most countries (19/24) also had sub-national differences in seasonality patterns, with strong differences in seasonality strength between regions. Seasonality patterns clustered into two macro-regions (West Africa and the Sahel vs. Eastern and Southern Africa), which were composed of sub-regional clusters with varying degrees of seasonality. Exploratory association analysis found most consistent and positive correlations between cholera seasonality and precipitation, and to a lesser extent with temperature and flooding.

**Interpretation:** Widespread cholera seasonality in SSA offers opportunities for intervention planning. Further studies are needed to study the association between cholera and climate.

**Funding:** The NASA Applied Sciences Program and the Bill and Melinda Gates Foundation.

**Research in Context:** *Evidence before this study:* Previous studies were searched using PubMed on Nov 15, 2021, without language or date restrictions using the search terms [“cholera*” AND “season*” AND (“Africa*” OR “Angola” OR “Burundi” OR “Benin” OR “Burkina Faso” OR “Botswana” OR “Central African Republic” OR “Côte d’Ivoire” OR “Cameroon” OR “Democratic Republic of the Congo” OR “Republic of Congo” OR “Djibouti” OR “Eritrea” OR “Ethiopia” OR “Gabon” OR “Ghana” OR “Guinea” OR “Gambia” OR “Guinea-Bissau” OR “Equatorial Guinea” OR “Kenya” OR “Liberia” OR “Madagascar” OR “Mali” OR “Mozambique” OR “Mauritania” OR “Malawi” OR “Namibia” OR “Niger” OR “Nigeria” OR “Rwanda” OR “Sudan” OR “Senegal” OR “Sierra Leone” OR “Somalia” OR “South Sudan” OR “São Tomé and Príncipe” OR “Swaziland” OR “Chad” OR “Togo” OR “Tanzania” OR “Uganda” OR “South Africa” OR “Zambia” OR “Zimbabwe”)]. Two additional known articles not identified by our PubMed search were added, one on the seasonality of cholera in Kenya and one on the epidemiology of cholera in West Africa. Studies were included if they focused on the epidemiology of *Vibrio cholerae* O1 or O139, covering one or multiple countries in sub-Saharan Africa (SSA) for at least two years with reported cases. We identified a total of 140 studies, of which 36 met our inclusion criteria. Of these 4 were reviews, 3 were regional or global studies, and 29 focused on specific countries. Local-level seasonality studies were identified in Burundi (1), Côte d’Ivoire (1), Cameroon (2), Democratic Republic of the Congo (3), Ghana (1), Guinea-Bissau (1), Kenya (1), Mozambique (3), Nigeria (1), Senegal (1), Somalia (1), South Sudan (1), Togo (1), Tanzania (4), Uganda (2), South Africa (1), and Zambia (4). These local studies mainly consisted of epidemiological descriptions of cholera incidence at either the national or first administrative unit scale, covering between 2 and 31 years of data. Most of these studies found seasonality in cholera with different patterns between countries. Two regional studies in West Africa found seasonal patterns in cholera incidence in the second part of the calendar year, with evidence for synchrony with rainfall patterns. Finally one global study between 1975 and 2005 found evidence for a latitudinal gradient of seasonality strength with weaker seasonality around the equator.

*Added value of this study:* Although most local-level and regional studies have found evidence for cholera seasonal patterns in SSA, gaps remain for certain countries and there currently lacks a systematic and continental-scale investigation. By leveraging a large database of cholera incidence we evaluate and characterize cholera seasonality at sub-national scale in 34 countries of SSA for which sufficient data was available. We show that cholera is seasonal in the majority of countries (24/34), with sub-national heterogeneity in most of them (19/24). Results enable the description of cholera seasonality at the continental level and the exploration of associations with hydro-climatic variables.

*Implications of all the available evidence:* This work establishes the extent and strength of cholera seasonality in SSA, providing a basis on which to ground strategic decision on large-scale intervention allocation as well as future work on the climatic and non-climatic drivers of cholera seasonality in SSA.

## Introduction

Despite being one of the oldest known infectious diseases, cholera, typically caused by toxigenic *Vibrio cholerae* bacteria of serogroup O1, still causes between 1 and 4 million cases per year ^1^. The majority of cholera cases reported to the World Health Organization (WHO) between 1996 and 2018, excluding the 2010 Haitian and 2017 Yemen epidemics, have occurred in sub-Saharan Africa (SSA), which also has the highest case fatality risks (2% in 2018) ^2^. An estimated 87 million people in SSA live in districts with high cholera incidence ^3^. Cholera transmission spans the endemic-epidemic spectrum across SSA, with large heterogeneity in transmission characteristics across time and space ^4,5^. Tailoring cholera prevention and control programs to local epidemiologic characteristics may be one efficient way to reach global targets for cholera control ^6^, though detailed systematic descriptions across broad geographies remain sparse.

Seasonality is one important aspect of cholera epidemiology and cholera exhibits strong seasonal patterns in countries on the Bay of Bengal, the ancestral homeland of cholera. The seasonal patterns of cholera in coastal and estuarine areas in this region have been linked in part to the ecology of *V. cholerae* in its natural brackish water habitats ^7,8^. Case studies from individual countries over short time-periods in SSA have demonstrated diverse seasonal patterns in cholera occurrence ^9–11^ though these fragmented descriptions have limited use in furthering our understanding of cholera dynamics and in global/regional public health planning. One of the major challenges hindering detailed large-scale descriptions of seasonality has been the lack of unified, fine-scale spatial and temporal resolution datasets on cholera occurrence.

Understanding seasonal variations in transmission of infectious diseases, like cholera, have direct implications for improving surveillance systems and tailoring control and elimination efforts ^12,13^. A better understanding of seasonality could allow for adaptive cholera surveillance and testing protocols, improvements in cholera risk assessments, improvements in planning cholera-prevention activities and could be used to help trigger local disease control activities. A detailed understanding of cholera seasonality can also enhance our ability to disentangle the links between cholera, climate and human behavior.

Here we aim to develop an ‘almanac’ of cholera seasonality across sub-Saharan Africa allowing for a sub-national understanding of how cholera risk varies throughout the year. We then use these results to explore the correlation between hydro-climatic variables and seasonality across the continent.

### Cholera data

The cholera incidence data used in these analyses come from a large database curated by Johns Hopkins University on behalf of the Global Taskforce on Cholera Control (GTFCC). Data consist of suspected and confirmed cholera case reports from various ministries of health, the World Health Organization (WHO), Médecins Sans Frontières, ProMED, ReliefWeb, scientific literature and publicly available epidemiologic reports. Suspected case definitions across counties, and time periods do vary, but are largely based on the recommended WHO and GTFCC case definitions^14^. Data resolution spanned multiple temporal (day to multi-year) and spatial (health zone/area to country) scales. Sub-monthly cholera incidence data were aggregated to the monthly time scale. Data were aggregated to both the first and second level administrative units ^15^ for separate analyses at these spatial scales. Details on data aggregation and availability per country are given in the Supplementary Material.

For our primary outcome variable, we defined a binary indicator of reported excess cholera occurrence, which equals one when the monthly cholera incidence rate for a given administrative unit exceeds the 2010-2016 estimated mean monthly incidence rate ^3^, or zero otherwise. In sensitivity analyses we explore two alternative definitions of cholera occurrence (i) ≥1 suspected case in a given month and (ii) ≥10 suspected cases in a given month. Of the 44 countries in SSA, we keep only those for which cholera was reported at a monthly or sub-monthly resolution data for at least 3 years of the study period. Based on this, we exclude 10 countries with insufficient data (Botswana, Republic of the Congo, Eritrea, Gabon, The Gambia, Equatorial Guinea, Mauritania, Rwanda, São Tomé and Príncipe, and Swaziland), comprising ∼1.2% of the SSA population, from the analysis due to lack of sub-yearly data.

### Model of cholera occurrence

We develop a model to infer the monthly relative risk of cholera occurrence at the administrative unit level accounting for inter-annual variability and spatial dependence. We build upon a Bayesian model commonly used for areal count data, which consists of a logistic regression with random effects for the month of the year (assumed to be temporally autocorrelated), year, as well as a combination of spatially correlated and uncorrelated random effects at the administrative unit level with an intrinsic conditional autoregressive prior ^16^. We fit the model to each country separately, and integrate multiple observations from different data sources covering the same month and administrative units by assuming that the combined reports of cholera excess occurrence follow a binomial distribution (e.g., if two data sources report excess cholera for a given administrative unit and month and a third does not, the data would be treated as two successes in three binomial trials). Observations covering multiple second-level administrative units and/or multiple months were included in the analysis by computing their corresponding probabilities of excess cholera occurrence (Supplementary Materials).

We considered a suite of different models that allowed for varying levels of flexibility in the seasonality of cholera within each country. First, we considered a model that has a single seasonal pattern per country, without assuming any particular shape (e.g., seasonality can be flat, unimodal, or multi-modal). We then expand this model to allow for two seasonal patterns within each country using a two-group mixture model with spatially auto-correlated grouping parameters. We also extended these models to include inference on the start of the “cholera year,” (e.g., the starting month that allowed for the best fit of both the seasonal and annual random effects, see Supplement for details), which does not necessarily align with the calendar year. We fit a total of five models for each country; no seasonality (‘null model’), a single seasonal pattern (‘base’), a single seasonal pattern with a distinct cholera year (‘offset’), two seasonal patterns (‘mixture model’), and a two seasonal patterns with a distinct cholera year (‘mixture offset’). Models were fitted using Markov Chain Monte Carlo methods with Rstan version 2.7. We ran 4 chains for each model with 1000 iterations (after 1000 warm-up) and assessed chain convergence using the Gelman-Rubin 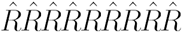 statistic^17^. For each country we selected the best fit model based on leave-one-out cross-validation^18^. Technical details are provided in the supplementary material.

### Seasonality Index

To characterize the strength of cholera seasonality, we computed a seasonality index for each administrative unit. This index is defined as the fraction of cholera risk that occurs during the cholera seasonality peak. The proportion was computed for each administrative unit as the sum of the mean predicted probabilities of occurrence within a 3 month window centered on the month of peak seasonality divided by the total sum of mean predicted probabilities.

### Spatial seasonality grouping

To characterize distinct seasonal cholera patterns across sub-Saharan Africa we used a soft-K-means clustering algorithm to infer groupings of administrative units with similarly shaped monthly relative risk curves ^19^. We implemented this algorithm in a Bayesian framework that accounts for spatial correlation between areas. We fit models with 2 to 10 seasonal groups (K) and compare model performance based on the estimated log-predictive density ^18^.

### Seasonality and climatology

We explored the association between the monthly relative risks of cholera with a suite of matched monthly hydroclimatic variables. These include 10km resolution estimates of 2m height air temperature generated by topographically downscaling the NASA Modern Era Reanalysis for Research and Applications v2 (MERRA-2) ^20^, monthly precipitation totals drawn from the Climate Hazards InfraRed Precipitation with Stations v2 (CHIRPSv2) ^21^ dataset, and 1/12º (∼10km) resolution estimates of monthly mean and maximum inundated area from the FloodScan multi-satellite product ^22^. Estimates of additional hydrological variables (soil moisture, runoff, and streamflow) were generated with an offline simulation of the Noah-Multi-Parameterization Land Surface Model ^23^ coupled to the Hydrological Modeling and Analysis Program (HyMAP) ^24^, using MERRA-2 and CHIRPSv2 as meteorological forcing. We quantified the associations between cholera monthly risk and hydroclimatic variables with Pearson correlation coefficients considering different lags between cholera and hydroclimatic variables (0-2 months).

Data used in these analyses were not identifiable and this work was deemed to be non-human subjects research by the Johns Hopkins IRB. Code, results and a minimal dataset are available at https://github.com/HopkinsIDD/cholera_seasonality_ssa/.

## Results

Our analysis dataset included 216’277 records of cholera incidence from 34 countries in sub-Saharan Africa between 1970 and 2021 spanning 2’529 distinct second-level administrative units, 427 first-level administrative units (97.3% of records since 2000, Supplementary Figure S1). Of these 200’844 (92.9%) were monthly or sub-monthly observations.

We found statistical support in 24 of 34 countries (84% of the SSA population) that excess cholera occurrence follows a distinct seasonal pattern (referred to throughout simply as ‘seasonality’, Supplementary Table 1, posterior predictive checks in Supplementary Figure S2). Countries for which a model with no seasonality was best supported were Burundi, Central African Republic, Djibouti, Ghana, Liberia, Madagascar, Namibia, Sudan, Senegal and Togo. Among countries with seasonal cholera, 79% (19/24) had evidence for within-country differences in seasonal cholera patterns (Supplementary Table S1).

Cholera seasonality showed distinct regional patterns (Figure 1, Supplementary Figures S3-S4 and S7-S8). Western Africa generally has higher excess cholera occurrence risk between July and October, in particular for inland countries across the Sahel, except for Mali. For example, the far north of Cameroon, had an 7.4-fold (95% CrI 3.0-18.7) increase in the odds of excess cholera in July compared to the average odds throughout the year. In Southern Africa and Mozambique, excess cholera risk generally peaked between December and April. Within Eastern Africa seasonal cholera patterns were more heterogeneous. Countries of the Great Lakes region as well as Angola had weak seasonality, peaking in the second half of the calendar year with Kenya and Tanzania having December to January peaks. South Sudan and Somalia had similar seasonality patterns with peak cholera risk between May and July. Finally, Ethiopia had a distinct North-South divide with the North peaking in August-September and the South having a weaker seasonal signal.

**Figure 1.**
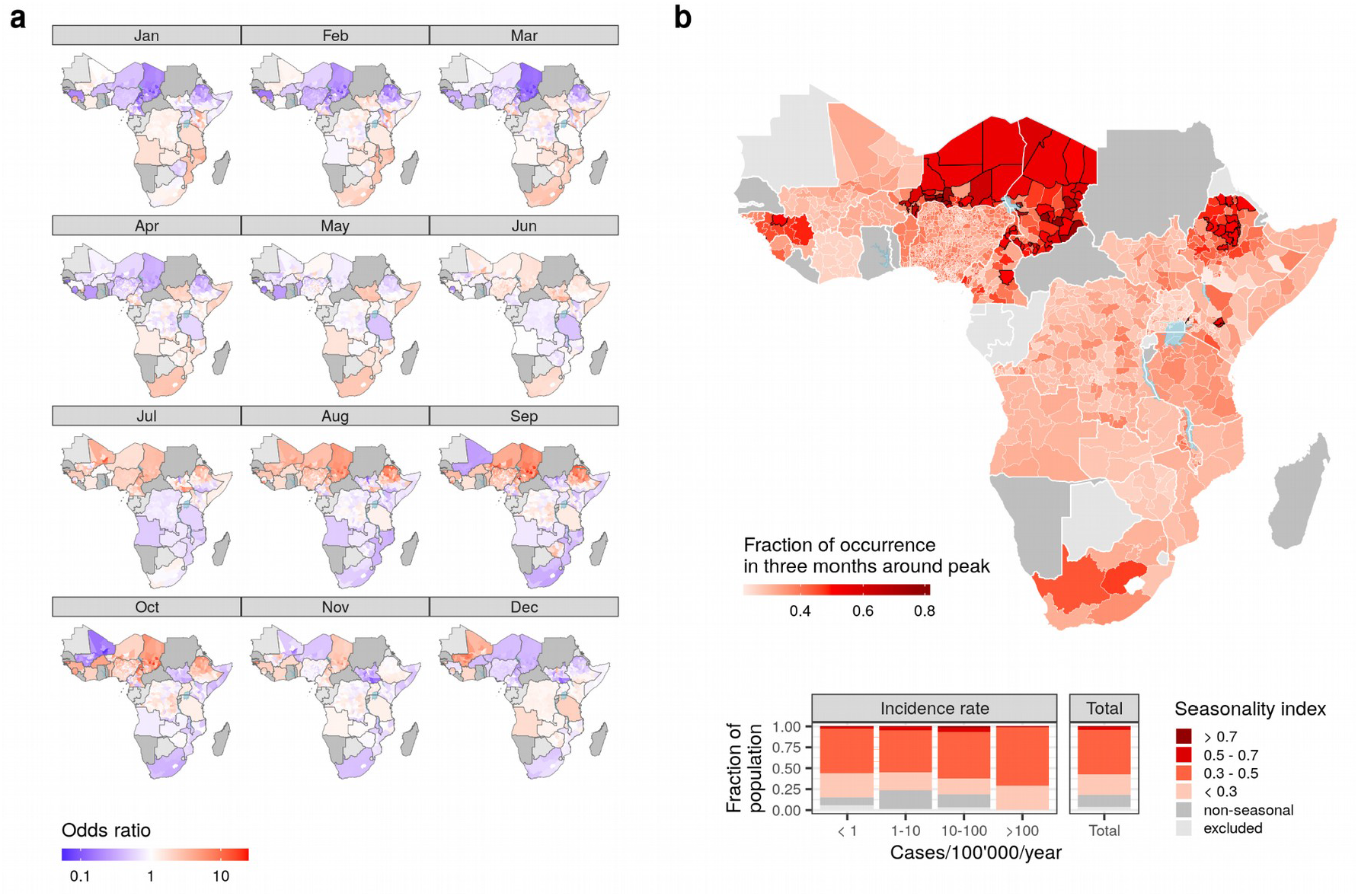
Cholera occurrence seasonality patterns and strength in sub-Saharan Africa. a) Estimates of monthly odds ratios of excess cholera occurrence defined as monthly incidence greater than the 2010-2016 baseline ^3^. Estimates are given for the best-performing model at the first or second administrative levels depending on data availability (Supplement), as opposed to countries for which the non-seasonal null model was selected (dark gray), and countries that were excluded from analyses due to lack of data (light gray). For illustrative purposes the odds ratios color scale was truncated at 20. Country-level time odds ratios time series are shown in Supplementary Figure S3. b) Strength of cholera seasonality quantified as the fraction of risk occurring in the three month window around the seasonal peak.

The strength of seasonality, quantified as the proportion of cholera risk in the three months around the peak, varied within and between countries (Figure 1b, uncertainty in Supplementary Figure S5). Seasonality was strongest in the Sahel and Northern Ethiopia. We subdivided administrative units in four categories of seasonality strength: those where cholera risk in the three months around the peak accounts for less than 30%, 30-50%, 50-70%, and more than 70% of the total excess cholera risk. The majority of the SSA population live in areas that report cholera 30-50% of the time within the three months period around the peak. The proportion of the population living in low seasonality areas (less than 30% of reporting in the three-month window) decreases as mean annual cholera incidence rates increase from less than 1 to 100 cases/100’000/year, and then decreases to similar levels as the total population for more than 100 cases/100’000/year. Overall, the strength of seasonality did not have a clear association with the population size, population density, area, nor mean annual incidence at the administrative unit level (Supplementary figure S6).

Cholera seasonality patterns across SSA clustered into two distinct macro-regions that partition sub-Saharan Africa into Western/Sahel and Eastern/Southern (Figure 2). On one hand, the macro-region consisting of Western Africa and the Sahel, including Northern Ethiopia and excluding Mali, had higher risk of excess cholera in the second half of the calendar year between July and October (labelled ‘Late peak’ in Figure 2). On the other hand, cholera tends to peak at the beginning of the year in Eastern and Southern Africa, and with less pronounced seasonality (labelled ‘Early peak’). Model comparison indicated statistical support for at least three groups, with similar levels of support for more than three clusters (Supplementary Figure S9). Increasing the number of clusters led to the partitioning of these two macro-regions in groups of administrative units with similar shapes, but different amplitude (Supplementary Figure S10). We here focus on clustering results with five groups composed of three degrees of seasonality amplitude in the Late peak macro-region, and two in the Early peak macro-region (Figure 2). Areas with a late seasonal peak and strongest amplitude were in the Northern Cameroon - Chad region, Northern Ethiopia, and Guinea. Late peak and moderate seasonality was mostly identified in Central Ethiopia and West Africa, except for areas in Niger and Nigeria which had weaker seasonality. The majority of the late peak macro-region was found to follow the higher amplitude pattern, with areas with lower amplitude mostly found in Central Africa including DRC, Uganda, and Tanzania; as well as Botswana and Western Zimbabwe. The repartition of people at risk of cholera among the three clusters (Figure 2a) echo results based on the seasonality index (Figure 1b). The majority of the SSA population live in administrative units in low amplitude seasonality, and the proportion of the population in clusters with stronger seasonality tends to increase with mean annual incidence rate although this stabilizes when incidence reaches 10 cases/100,000/year.

**Figure 2:**
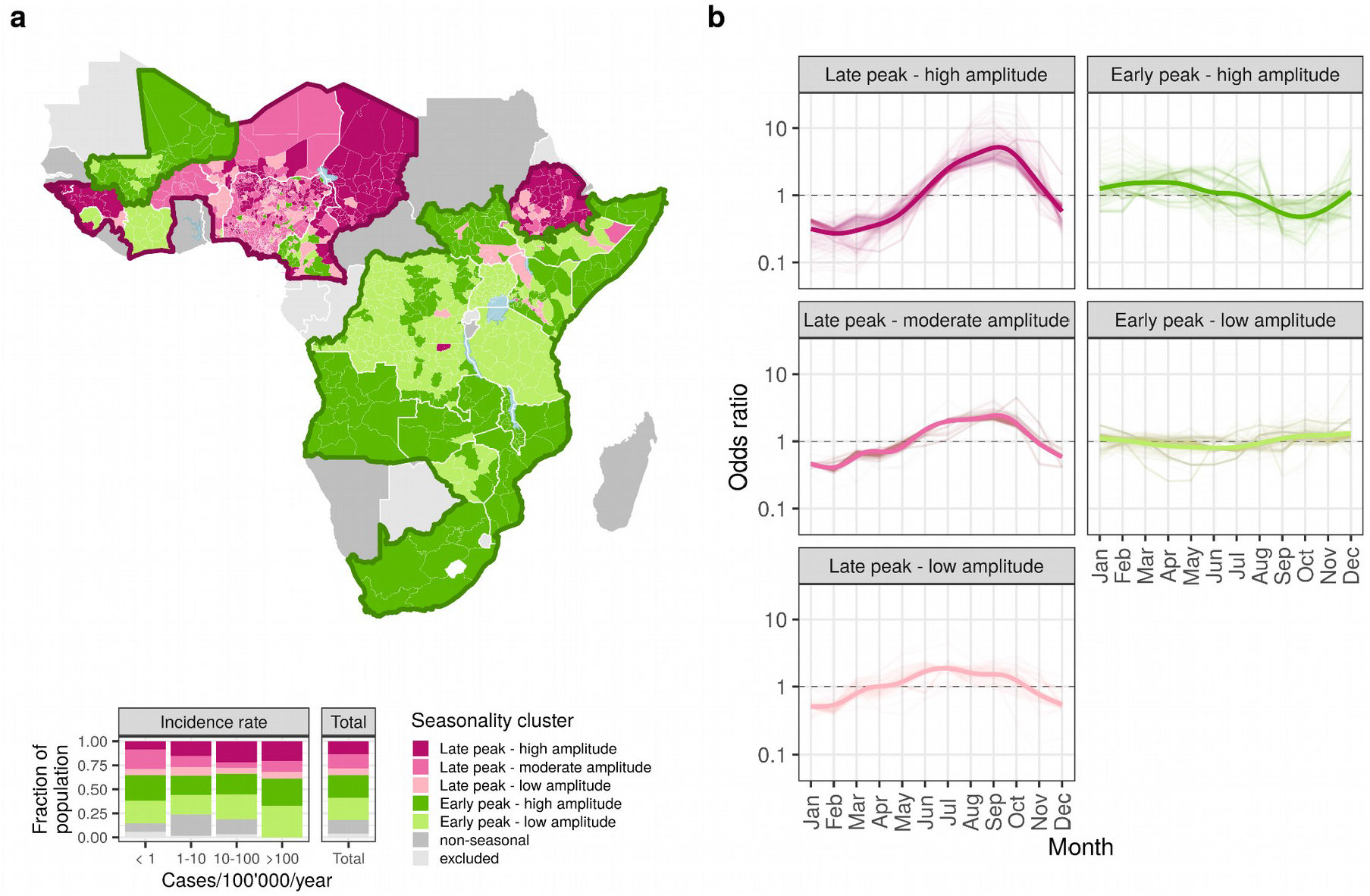
Seasonality grouping in sub-Saharan Africa. a) Map of cholera seasonality clusters (top) and fraction of population in each cluster for different categories of mean annual cholera incidence (bottom). Clustering results are shown for models with two (think borders) and five (color fill) clusters, along with countries for which seasonality was not retained (dark gray) and excluded because of lack of data (light gray). Marco-regions of the two-cluster model were outlined using the convex hull of the corresponding administrative units. b) Seasonality of cholera excess risk in each cluster for each administrative unit (light lines), and overall trend (full lines) estimated by a GAM of odds ratio as a function of the month of the year.

We explored the correlation between the monthly odds ratios of excess cholera and mean monthly fraction of area flooded, mean monthly air temperature, and cumulative monthly precipitation with lags of 0,1 and 2 months. We found large differences in the direction and strength of these relationships across SSA (Figure 3, Supplementary Figure S11). We find that correlation between monthly excess cholera risk and flooding was generally weak in most countries; and when present, could go in both directions. We find that the spatial extent of areas where excess cholera and mean temperature had significant correlation was limited, with both positive (Tanzania, Northern and Eastern DRC) and negative (Eastern Ethiopia, Northern Ivory Coast, Southern Chad) associations, although the spatial extent and strength of correlation increased with lags, particularly in southern Africa. Excess cholera was most consistently associated with rainfall, showing mostly strong positive correlation across SSA (69.6% of admin units in countries with seasonality in SSA had correlation ≥0.5). We identified two geographic areas with large positive correlation around lake Chad (Niger, Nigeria, Northern Cameroon and Chad), Eastern Ethiopia, the central part of Eastern Africa (Botswana, Malawi, Mozambique, Zambia and Zimbabwe). Associations with other hydro-climatic variables and lags are given in Supplementary Figure S12.

**Figure 3.**
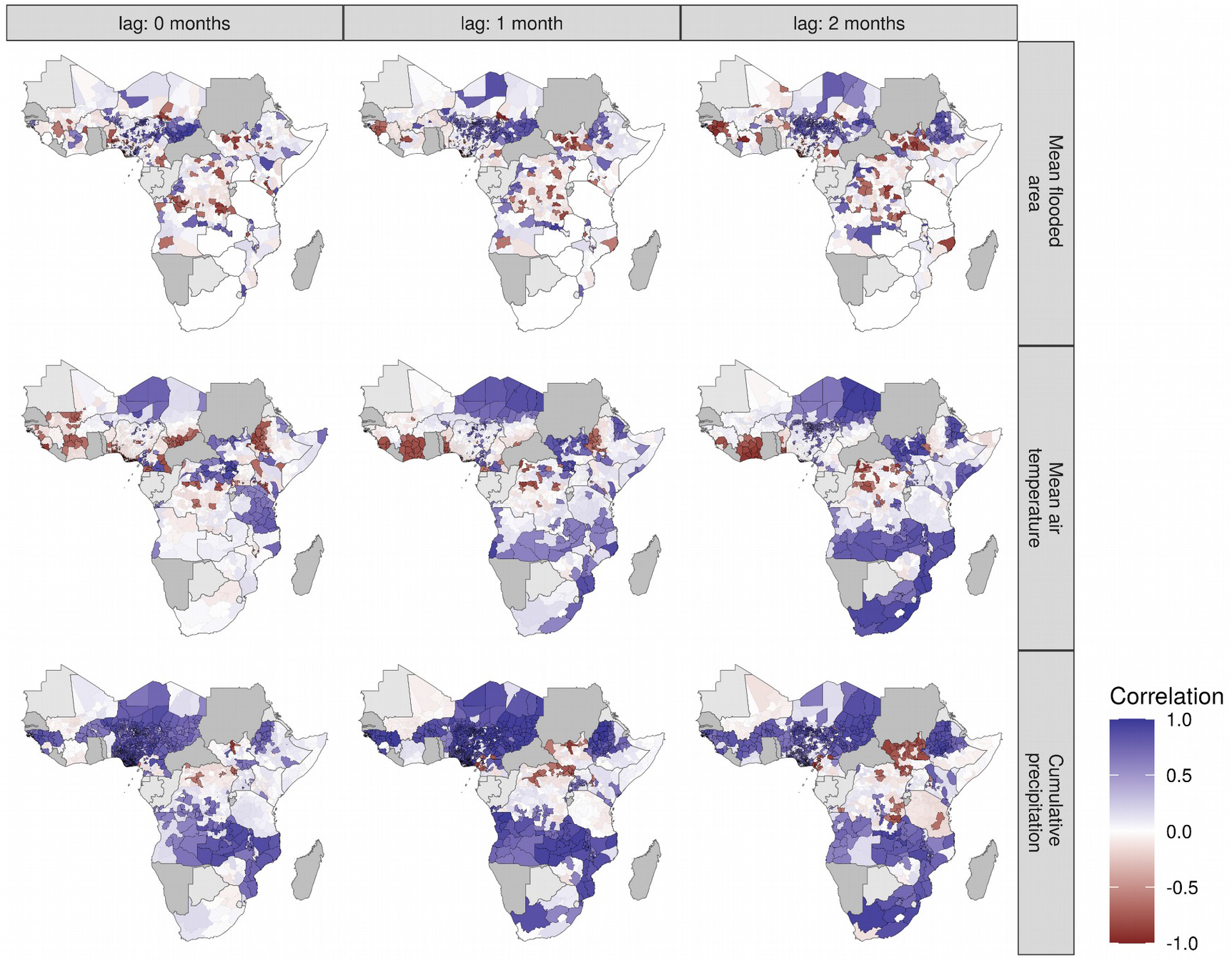
Excess cholera seasonality and climatology. Maps show the Spearman correlation between excess cholera odds ratios and the mean monthly values of hydro-climatic variables at 0, 1, and 2 month lags. Hydro-climatic variables include the mean monthly flooded area, the mean monthly air temperature, and the monthly cumulative precipitation. Correlation is shown for significant coefficients (p-value < 0.05, full color, black border), with non-significant values (p-value > 0.05) given for indication (transparent, no border). Associations with other hydro-climatic variables given in Supplementary Figure S8.

## Discussion

We found that more than around 85% of people in sub-Saharan Africa live in locations with significant seasonal variations in excess cholera occurrence, although patterns of seasonality vary between and within countries. Seasonality was stronger in West Africa than Eastern and Southern Africa, with the largest seasonality strength in the Sahel. These differences mapped to macro-regions with distinct seasonality patterns, within which there was varying degrees of seasonality amplitude. Seasonality patterns correlated most consistently and strongly with mean monthly precipitation, with areas of high correlation in the countries bordering lake Chad as well as South-Eastern Africa. Taken together these findings suggest that cholera can be considered a seasonal disease in most sub-Saharan countries having mixed associations with hydro-climatic factors.

The results of this study support the characterization of cholera epidemiology from local analysis across countries in sub-Saharan Africa ^9,25–29^. The identification of areas with synchronous patterns of cholera occurrence had been highlighted before in Western Africa ^30^, as well as in the Great Lakes region ^11^. By combining data at the regional level we show that cholera occurrence seasonality is aligned at larger spatial scales with two main synchronous regions, one composed of West Africa and Northern East-Africa, and one composed of East and Southern Africa. We note however that the alignment of our estimates of seasonality does not imply synchrony of outbreaks due to their multiple drivers including human introductions of *V. cholerae*, changes in vulnerability, or changes in the immune landscape, and extrinsic factors like natural disasters in explaining the strong inter-annual variability of cholera epidemics ^4,5^. Nevertheless the availability of large-scale estimates of seasonality provides an opportunity to formulate hypotheses on driving mechanisms in complement to local-scale studies, including pathogen introductions, variations in transmission, changes in socio-behavioral factors, and seasonal patterns of water use.

The correlations between hydro-climatic variables and the seasonality of cholera occurrence contribute to the complex picture of cholera and climate in SSA. The lack of widespread positive correlation with mean monthly temperature, especially in all coastal areas and around large inland lakes, is further evidence against the predominant role of *V. cholerae*’s environmental dynamics in driving cholera outbreaks in the region ^5^, which would imply spatial synchrony between seasonality and temperature (the Moran effect). On the other hand, the presence of large areas of positive correlation with cumulative monthly precipitation support previous country-specific findings across the sub-continent ^10,11,25,30^. These correlations hint towards the role of rainfall-driven fecal contamination of water sources in cholera occurrence, as it has been suggested for different settings ^31–33^. However our estimates are only correlations between the monthly odds ratio of excess cholera and mean monthly hydro-climatic variables, which should not be interpreted causally. The relationship between seasonal cholera occurrence and climatic drivers has further been suggested by the indirect role of El Niño Southern Oscillation: the spatial distribution of cases in SSA from Western to Eastern Africa between El Niño and non-El Niño years, through differential sensitivity to climate anomalies ^34^. In the absence of a conceptual framework of the relationship between cholera and climate, further investigation would benefit from explicit hypotheses on the role of hydro-climatic variables on cholera occurrence, as well accounting for possible nonlinearities in their effects and interactions with social factors ^35^.

These results come with several limitations. Data used to define cholera occurrence consisted of suspected cholera cases due to the small proportion of suspected cholera cases that are laboratory confirmed across SSA and the world. The definition of suspected cholera cases may vary between settings but typically follow the WHO and GTFCC recommended case definitions. As such cholera seasonality estimates in this work may be confounded by the seasonality of other aetiologies of watery diarrhea. Cholera incidence estimates used to define excess occurrence, i.e. months with cases above the mean monthly incidence, correspond to the period 2010-2016, whereas most of the cholera data used in this analysis spanned the years 2000 to 2021 which may alter the classification of cholera presence/absence. Seasonality estimates and the main results of the analysis remained valid in sensitivity analysis using alternative definitions of cholera occurrence (more than 1 and more than 10 suspected cases, Supplementary Figure S13). Finally, an assumption in the analysis is that seasonality did not change during the study period.

Despite these limitations the work has public health implications from the perspective of the GTFCC’s 2030 Cholera Roadmap ^36^ The presence and strength of seasonality in cholera occurrence provides an opportunity for timing preventive interventions and preparedness activities for outbreak response in periods of low odds of occurrence. Country and sub-national level estimates of seasonality as produced in this study can serve as a basis for these efforts, as well as for identifying outbreaks that occur prior to the expected season start.

## Data Availability

Code, results and a minimal dataset are available at https://github.com/HopkinsIDD/cholera_seasonality_ssa/

## Acknowledgements

The authors thank Josh Kaminsky, Maya Demby and Qulu Zheng for their assistance with the cholera database. We thank all of the Ministries of Health, WHO offices, and the UNICEF WCAR and ESAR cholera platforms for contributing data to the database. We thank the GTFCC for helpful discussion, and the Johns Hopkins Infectious Disease Dynamics group for feedback on study design and analysis.

## Author Contributions

JPS: conceptualisation; data curation; formal analysis; methodology; software; visualisation

writing – original draft; writing– review & editing

JL: funding acquisition; validation; writing– review & editing

EL: data curation; writing– review & editing

FJL: validation, writing– review & editing

EBM: writing– review & editing

FF: writing– review & editing

JPL: writing– review & editing

BZ: funding acquisition; resources; validation; writing– review & editing

ASA: conceptualisation; funding acquisition; methodology; project administration; validation; supervision; writing – original draft; writing– review & editing

