## Supplementary Material for "The seasonality of cholera in sub-Saharan Africa"

#### Contents

**S1 Cholera data and processing** S1
S1.1 Spatial matching and aggregation to GADM Units . . . . . S1
S1.2 Temporal aggregation . . . . . S2
S1.3 Selection of GADM level for analysis . . . . . S2

**S2 Seasonality model description** S3
S2.1 Base model . . . . . S3
S2.2 Offset model . . . . . S4
S2.3 Mixture model . . . . . S5
S2.4 Priors . . . . . S5

**S3 Model comparison** S6

**S4 Posterior predictive checks** S9

**S5 Additiona results** S10

**S6 Seasoanlity grouping** S14

**S7 Correlation with hydro-climatic variables** S16

**S8 Alternative cholera occurrence definitions** S18

#### S1 Cholera data and processing

Cholera incidence data consisted of reports from various sources (ministries of health, WHO, Médecins Sans Frontières, ProMED, ReliefWeb, scientific literature and publicly available epidemiologic reports) with different spatial and temporal resolutions. We define as a location-period (LP) a set of spatial polygons corresponding to the areas where cholera was reported, and a temporal window covered by a the incidence report (for instance a given first level administrative unit during a specific epidemiological week). LPs of cholera reports contained in the database were aggregated in space and time to obtain the analysis datasets (first and second administrative levels at the monthly temporal resolution) on which models were run.

##### S1.1 Spatial matching and aggregation to GADM Units

Our spatial reference for analysis is the GADM Global Administrative Areas dataset (Hijmans, Garcia, and Wieczorek 2010). We first match LPs to GADM units at the country, first and second administrative levels.

An LP was defined to match a GADM unit if: 1) the centroids of the two where mutually contained withing each other, 2) the LP's area within 90% and 110% the size of the GADM unit's area. For the set of LPs that where within a given GADM units but for which the area was smaller than the 90% threshold we compared the union of their areas to that of the GADM unit within each data source, and flagged whether the union of areas was below, within, or above the 90-110% range used for determining matching. This flag was then used in temporal aggregation.

#### S1.2 Temporal aggregation

Analysis were run at the monthly time scale, we thus aggregated all sub-monthly observations in each LP to the monthly resolution. These monthly aggregates where then used to define cholera excess occurrence by comparing monthly incidence to the mean monthly incidence estimates extracted from Lessler et al. (2018). Observations of length longer than 1 month were not modified and were compared to the mean incidence with the exposure period corresponding. In cases where the union of areas of matching LPs flagged during spatial aggregation we applied the following rule: 1) if the union of areas was below the 90% threshold we set cholera occurrence to 1 if the monthly incidence was larger than the mean monthly incidence, and dropped the observation if it was smaller; 2) if the union of areas was larger than the 110% threshold we set cholera occurrence to 0 if the monthly incidence was bellow the mean monthly estimate, and dropped the observation if it was larger.

#### S1.3 Selection of GADM level for analysis

We ran all models both at first and second administrative levels. For analysis we selected the GADM level for each country based on data availability. In each country, we computed the yearly fraction of observed administrative 2 level units for observations of length 1 to 3 months. We select the second administrative level if the mean fraction across years was larger than 10% and the maximum larger than 40%. If these conditions were not met we selected the first administrative level.

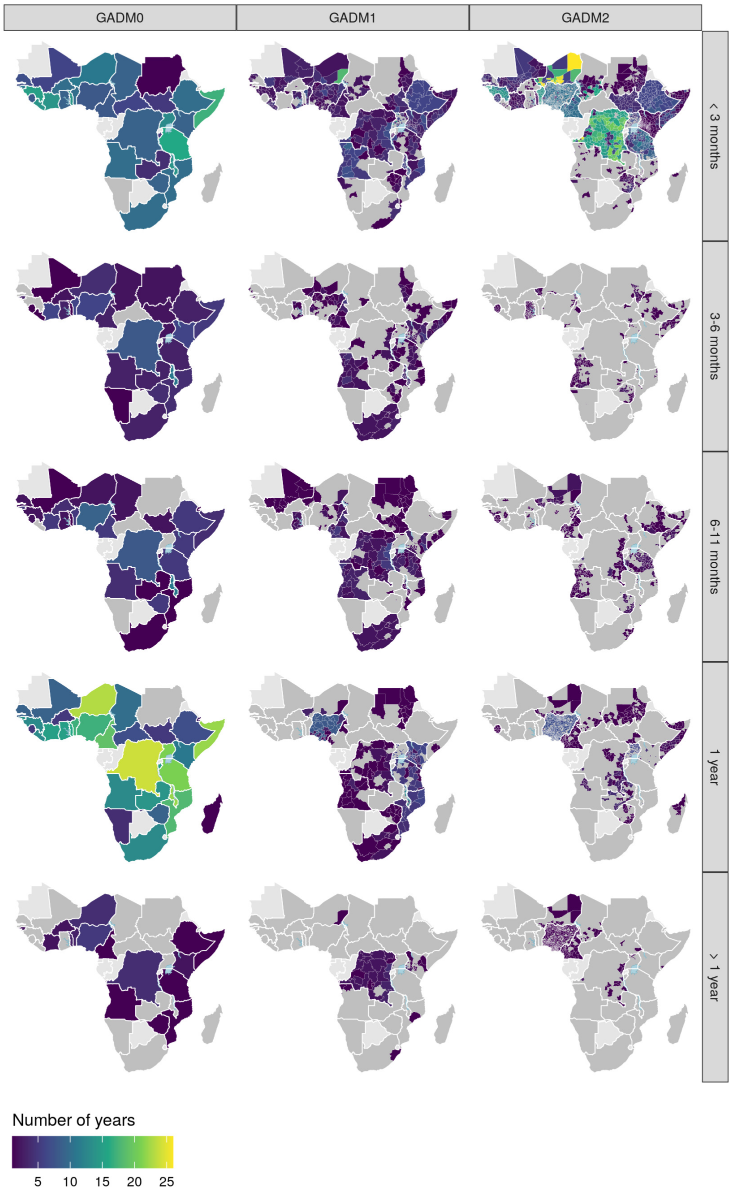


Figure S1: Map of cholera incidence data availability in sub-Saharan Africa. Availability is expressed in terms of the number of distinct years for which at least one observation is available, partitioned by GADM administrative level and obervevation length. Light gray indicate countries that were excluded due to lack of data.

### S2 Seasonality model description

#### S2.1 Base model

Cholera occurrence is modeled as binary observation process at a monthly temporal resolution. The analysis is done on a per-country basis taking the administrative subdivisions as the spatial units of the disease process. The aim is to draw on information on all administrative levels by exploiting the nested structure of the administrative units for instance districts within provinces within a country.

The data consists observations of cholera occurrence per month. Assuming that the administrative level 2 is the lowest administrative level at which cholera is observed, we assume that the set of $n_{i,m,y}$ monthly observations for month $m$ in administrative level 2 unit $i$ of year $y$, $Y_{i,m,y}$, result from a binomial distribution:

$$Y_{i,m,y} \sim \text{Binomial}(n_{i,m,y}, p_{i,m,y}),$$

where $p_{i,m,y}$ is the combined probability of there being cholera and observing it (the process and measurement models are combined). The monthly probabilities are assumed to follow the BYM model:

$$\text{logit}(p_{i,m,y}) = \beta_m + \eta_y + \theta_i + \phi_i,$$

where $\beta_m$ is the monthly country-level cholera probability of month $m$, $\eta_y$ is a year-specific random effect, and $\theta_i$ and $\phi_i$ are the non-spatial and spatial random effects. We assume that the $\beta$s follow a 1-dimensional cyclic random walk, i.e. $\beta_m - \beta_{m-1} \sim \mathcal{N}(0, \tau_\beta^{-1})\ \forall m \in [2, 12]$ and $\beta_1 - \beta_{12} \sim \mathcal{N}(0, \tau_\beta^{-1})$ to impose the cyclic pattern. To allow the identifiability of the model we further impose $\sum_{m=1}^{12} \beta_m = 0$. We assume that the spatial random effects follow the ICAR model, and use the reparameterized version of the BYM model (Riebler et al. 2016):

$$\theta_i + \phi_i = \epsilon_i = \frac{1}{\sqrt{\tau}} \left( \sqrt{1-\rho}\theta_i^* + \sqrt{\frac{\rho}{s}}\phi_i \right),$$

where $\tau$ is the overall marginal precision (i.e. the total residual relative risk), $\theta^* \sim \mathcal{N}(0, 1)$, $\phi \sim ICAR$, $s$ is a scaling factor that depends on the adjacency matrix of the neighborhood network to ensure $Var(\phi^*) \approx 1$, and $\rho \in [0, 1]$ indicates whether the marginal variance comes from spatial or non-spatial random effects.

At the upper administrative level the observation process at the admin unit level 1 $j$ is given by the probabilities of cholera reporting in all areas at the level 2 contained in it, $i \sim j$, as:

$$Y_{j,m,y} \sim \text{Binomial}(n_{j,m,y}, p_{j,m,y}) p_{j,m,y} = 1 - \prod_{i \sim j} 1 - p_{i,m,y}.$$

The country-level monthly observations follow the same observation process with the product over all spatial subunits.

We also account for multi-month observations. The probability of observing cholera in admin unit $i$ in year $y$ for period $T_n = \{m_1, \ldots, m_n\}$ composed of $n$ months is given by:

$$p_{i,T_n,y} = 1 - \prod_{m=m_1}^{m_n} 1 - p_{i,m,y}.$$

Similarly multi-month observations at any upper admin level $j$ is obtained by multiplying the individual probabilities:

$$p_{j,T_n,y} = 1 - \prod_{i \sim j} \prod_{m=m_1}^{m_n} 1 - p_{i,m,y}.$$

#### S2.2 Offset model

The base model considers yearly random effects based on the calendar year, which may be unrealistic in some situations. For instance if cholera has a single peak in December/January calendar-based random effects will apply only to the first "half" the length of the epidemic with the second half falling in the following calendar year.

To account for this fact we introduce and offset in the assignment of the yearly random effect based on the month in order to align the yearly random effects to "cholera" years by introducing a latent discrete variable representing the offset. The probability of occurrence becomes:

$$logit(p_{i,j,m}) = \beta_m + \eta_{f(yr,m,z)} + \epsilon_i,$$

where $f(yr, m, z)$ gives the year corresponding to the month accounting for the offset.

$$f(yr,m,z) = \begin{cases} yr & \text{if} \quad m+z \leq 12 \\ yr+1 & \text{if} \quad m+z > 12 \end{cases}, \quad z \in [0, 11].$$

For the purpose of sampling $z$ can be marginalized out of the likelihood as:

$$\begin{aligned} p(Y|\Theta) &= \sum_{z=0}^{11} p(Y|\Theta, z)p(z), \\ &= \sum_{z=0}^{12} \text{Uniform}(z|0,11) \prod_{j=1}^{n_{obs}} \text{Binomial}(Y_j|n_j, p_{j,z}) \end{aligned}$$

where $\Theta$ is the vector of all parameters defining the occurrence probabilities $(\beta, \eta, \rho, \tau, \phi, \theta, \xi)$ subscript $j$ indicates the observation (combination of admin level, month and year).

And the log-likelihood as:

$$\log(p(Y|\Theta)) = \log\left\{\sum_{z=0}^{11} \exp(ll_z)\right\},$$

where $ll_z$ are the individual sum of marginal log-likelihoods and the log of the prior on $z$:

$$ll_z = \log(p(Y|\Theta, z)p(z)) = \log(\text{Uniform}(z|0,12)) + \sum_{j=1}^{n_{obs}} \log(\text{Binomial}(Y_j|n_j, p_{j,z})).$$

#### S2.3 Mixture model

Assuming the possible coexistence of two distinct groups of seasonalities within the same country, a latent class model for the probabilities of observing cholera can be written in terms of a mixture of two different vectors of seasonality coefficients $\beta^{g_1, g_1}$:

$$logit(p_{i,j,m}) = \lambda_i \beta_m^{g_1} + (1-\lambda_i)\beta_m^{g_2} + \eta_{f(yr,m,z)} + \epsilon_i,$$

with $\lambda_i \in (0, 1)$ representing the mixture probabilities, with priors $\lambda \sim beta(0.5, 0.5)$. To mitigate label switching issues we impose the constraints $\beta^{g_2}[1] > \beta^{g_1}[1]$ and $\beta^{g_2}[7] < \beta^{g_1}[7]$.

#### S2.4 Priors

We use the following priors:

$$\begin{aligned} \tau_\beta &\sim \mathcal{N}(0, 2.5), \\ \eta_t &\sim \mathcal{N}(0, \sigma_\eta), \\ \sigma_\eta &\sim \mathcal{N}(0, 2.5). \end{aligned}$$

Penalized-complexity (PC) priors are chosen for on $\tau$ and $\phi$ following (Riebler et al. 2016): $\pi(\tau) = \frac{\theta}{2}\tau^{-3/2}\exp\left(-\theta\tau^{-1/2}\right)$, with $\theta = -\log(\alpha_\tau)/U_\tau$ where $P\left(\frac{1}{\sqrt{\tau}} > U_\tau\right) = \alpha_\tau$ with $\alpha_\tau, U_\tau$ set to represent our belief in the importance of marginal variance. Similarly a PC prior for $\rho$ can be formulated in terms of the probability $P(\rho < U_\rho) = \alpha_\rho$, which does not have a closed-form expression but for which tabulated values can be computed.

### S3 Model comparison

Table S1: Model fit results and model selection. ELPD diff: difference in estimated log predictive density with respect to best-fitting model. SE: standard error of the ELPD difference. LOO-IC: Leave-one-out cross-validation estimate Information Criterion. Rank: model rank. Weight: Evidence weight in favor of the model.

| model | ELPD diff | SE | LOO-IC | rank | weight |
|---|---|---|---|---|---|
| **Angola** | | | | | |
| offset | - | - | 1618.8 | 1 | 1.00 |
| mixture_offset | -34.3 | 6.5 | 1687.3 | 2 | 0.00 |
| mixture | -95.3 | 14.5 | 1809.4 | 3 | 0.00 |
| base | -97.1 | 14.3 | 1813.1 | 4 | 0.00 |
| null | -114.7 | 16.3 | 1848.1 | 5 | 0.00 |
| **Benin** | | | | | |
| mixture | - | - | 1082.2 | 1 | 0.88 |
| base | -2.0 | 2.1 | 1086.1 | 2 | 0.12 |
| null | -27.6 | 7.8 | 1137.5 | 3 | 0.00 |
| mixture_offset | -32.5 | 6.4 | 1147.2 | 4 | 0.00 |
| offset | -94.6 | 9.3 | 1271.3 | 5 | 0.00 |
| **Botswana** | | | | | |
| offset | - | - | 29.9 | 1 | 0.85 |
| mixture_offset | -2.7 | 0.6 | 35.3 | 2 | 0.06 |
| mixture | -3.1 | 1.4 | 36.1 | 3 | 0.04 |
| base | -3.2 | 1.4 | 36.3 | 4 | 0.03 |
| null | -3.7 | 1.3 | 37.4 | 5 | 0.02 |
| **Burkina Faso** | | | | | |
| base | - | - | 29.7 | 1 | 0.60 |
| mixture | -0.7 | 0.3 | 31.0 | 2 | 0.31 |
| null | -2.3 | 1.0 | 34.3 | 3 | 0.06 |
| mixture_offset | -3.2 | 1.5 | 36.1 | 4 | 0.03 |
| offset | -9.6 | 3.3 | 48.9 | 5 | 0.00 |
| **Burundi** | | | | | |
| mixture_offset | - | - | 175.0 | 1 | 0.42 |
| null | -0.2 | 1.2 | 175.3 | 2 | 0.36 |
| mixture | -0.9 | 1.3 | 176.8 | 3 | 0.17 |
| base | -2.1 | 1.5 | 179.2 | 4 | 0.05 |
| offset | -17.0 | 2.8 | 209.1 | 5 | 0.00 |
| **Côte d'Ivoire** | | | | | |
| offset | - | - | 130.5 | 1 | 0.98 |
| mixture_offset | -3.7 | 2.2 | 137.9 | 2 | 0.02 |
| mixture | -12.4 | 3.6 | 155.3 | 3 | 0.00 |
| base | -12.5 | 3.7 | 155.5 | 4 | 0.00 |
| null | -14.3 | 3.7 | 159.1 | 5 | 0.00 |
| **Cameroon** | | | | | |
| mixture | - | - | 2367.0 | 1 | 1.00 |
| mixture_offset | -17.5 | 14.9 | 2402.0 | 2 | 0.00 |
| base | -60.9 | 10.7 | 2488.7 | 3 | 0.00 |
| null | -86.7 | 14.5 | 2540.5 | 4 | 0.00 |
| offset | -303.7 | 20.5 | 2974.5 | 5 | 0.00 |
| **Central African Republic** | | | | | |
| mixture_offset | - | - | 57.0 | 1 | 0.77 |
| base | -1.7 | 2.0 | 60.3 | 2 | 0.15 |
| mixture | -2.9 | 2.7 | 62.8 | 3 | 0.04 |
| offset | -3.0 | 1.5 | 62.9 | 4 | 0.04 |
| null | -5.1 | 2.6 | 67.2 | 5 | 0.00 |
| **Congo** | | | | | |
| mixture_offset | - | - | 53.4 | 1 | 0.31 |
| null | -0.2 | 1.0 | 53.7 | 2 | 0.26 |
| mixture | -0.4 | 1.1 | 54.3 | 3 | 0.20 |
| base | -0.5 | 1.4 | 54.5 | 4 | 0.18 |
| offset | -1.9 | 1.7 | 57.2 | 5 | 0.05 |
| **Democratic Republic of the Congo** | | | | | |
| mixture | - | - | 16416.5 | 1 | 1.00 |
| base | -110.2 | 16.4 | 16636.9 | 2 | 0.00 |
| null | -131.4 | 18.7 | 16679.4 | 3 | 0.00 |
| mixture_offset | -188.2 | 27.2 | 16792.9 | 4 | 0.00 |
| offset | -211.6 | 44.5 | 16839.7 | 5 | 0.00 |
| **Djibouti** | | | | | |
| null | - | - | 25.6 | 1 | 0.28 |
| offset | -0.3 | 1.0 | 26.2 | 2 | 0.22 |
| mixture | -0.4 | 0.3 | 26.5 | 3 | 0.19 |
| mixture_offset | -0.4 | 0.7 | 26.5 | 4 | 0.19 |
| base | -0.8 | 0.4 | 27.3 | 5 | 0.12 |
| **Ethiopia** | | | | | |
| mixture_offset | - | - | 3021.5 | 1 | 1.00 |
| mixture | -116.5 | 15.9 | 3254.4 | 2 | 0.00 |
| base | -161.1 | 18.0 | 3343.7 | 3 | 0.00 |
| null | -261.4 | 22.2 | 3544.3 | 4 | 0.00 |
| offset | -412.8 | 22.0 | 3847.1 | 5 | 0.00 |

*(Continued on Next Page...)*

Table S1: *(continued)*

| model | ELPD diff | SE | LOO-IC | rank | weight |
|---|---|---|---|---|---|
| **Ghana** | | | | | |
| mixture | - | - | 305.4 | 1 | 0.30 |
| mixture_offset | -0.0 | 8.4 | 305.4 | 2 | 0.29 |
| null | -0.1 | 1.4 | 305.6 | 3 | 0.27 |
| base | -0.8 | 0.9 | 306.9 | 4 | 0.14 |
| offset | -21.9 | 7.6 | 349.2 | 5 | 0.00 |
| **Guinea** | | | | | |
| mixture | - | - | 664.8 | 1 | 1.00 |
| base | -16.3 | 5.2 | 697.4 | 2 | 0.00 |
| offset | -50.5 | 8.6 | 765.8 | 3 | 0.00 |
| null | -69.2 | 12.2 | 803.1 | 4 | 0.00 |
| **Guinea-Bissau** | | | | | |
| base | - | - | 654.7 | 1 | 0.92 |
| mixture | -2.4 | 2.8 | 659.6 | 2 | 0.08 |
| null | -84.2 | 16.3 | 823.1 | 3 | 0.00 |
| offset | -358.9 | 31.9 | 1372.6 | 4 | 0.00 |
| mixture_offset | -1421.0 | 249.1 | 3496.8 | 5 | 0.00 |
| **Kenya** | | | | | |
| mixture | - | - | 1997.1 | 1 | 1.00 |
| null | -39.8 | 9.5 | 2076.7 | 2 | 0.00 |
| base | -42.5 | 8.7 | 2082.2 | 3 | 0.00 |
| mixture_offset | -1695.2 | 161.0 | 5387.5 | 4 | 0.00 |
| **Liberia** | | | | | |
| mixture_offset | - | - | 359.9 | 1 | 0.99 |
| mixture | -5.4 | 6.5 | 370.7 | 2 | 0.00 |
| base | -6.1 | 6.7 | 372.0 | 3 | 0.00 |
| null | -6.5 | 6.5 | 372.9 | 4 | 0.00 |
| offset | -68.0 | 9.3 | 495.9 | 5 | 0.00 |
| **Madagascar** | | | | | |
| base | - | - | 1.0 | 1 | 0.21 |
| null | -0.0 | - | 1.0 | 2 | 0.20 |
| mixture | -0.0 | - | 1.0 | 3 | 0.20 |
| mixture_offset | -0.1 | - | 1.1 | 4 | 0.20 |
| offset | -0.1 | 0.1 | 1.2 | 5 | 0.19 |
| **Malawi** | | | | | |
| mixture | - | - | 1503.4 | 1 | 1.00 |
| offset | -6.9 | 12.4 | 1517.3 | 2 | 0.00 |
| base | -19.5 | 5.5 | 1542.3 | 3 | 0.00 |
| null | -52.5 | 9.8 | 1608.4 | 4 | 0.00 |
| mixture_offset | -56.6 | 24.9 | 1616.6 | 5 | 0.00 |
| **Mali** | | | | | |
| mixture | - | - | 393.0 | 1 | 1.00 |
| base | -10.9 | 4.1 | 414.8 | 2 | 0.00 |
| null | -38.1 | 11.7 | 469.1 | 3 | 0.00 |
| mixture_offset | -44.0 | 9.4 | 481.0 | 4 | 0.00 |
| offset | -211.8 | 11.6 | 816.6 | 5 | 0.00 |
| **Mauritania** | | | | | |
| mixture | - | - | 195.0 | 1 | 1.00 |
| base | -6.0 | 2.6 | 207.0 | 2 | 0.00 |
| mixture_offset | -17.7 | 3.7 | 230.4 | 3 | 0.00 |
| null | -35.3 | 6.1 | 265.7 | 4 | 0.00 |
| offset | -84.1 | 4.6 | 363.3 | 5 | 0.00 |
| **Mozambique** | | | | | |
| mixture | - | - | 222.5 | 1 | 0.93 |
| base | -2.5 | 1.0 | 227.6 | 2 | 0.07 |
| null | -8.6 | 3.2 | 239.7 | 3 | 0.00 |
| offset | -44.7 | 9.6 | 311.9 | 4 | 0.00 |
| mixture_offset | -764.2 | 154.7 | 1750.9 | 5 | 0.00 |
| **Namibia** | | | | | |
| base | - | - | 20.2 | 1 | 0.35 |
| null | -0.1 | 0.3 | 20.3 | 2 | 0.34 |
| mixture | -0.1 | 0.2 | 20.4 | 3 | 0.31 |
| mixture_offset | -7.9 | 3.3 | 36.0 | 4 | 0.00 |
| offset | -13.9 | 4.1 | 47.9 | 5 | 0.00 |
| **Niger** | | | | | |
| mixture | - | - | 2513.2 | 1 | 1.00 |
| base | -25.8 | 8.2 | 2564.8 | 2 | 0.00 |
| mixture_offset | -27.9 | 18.2 | 2569.1 | 3 | 0.00 |
| offset | -137.9 | 22.5 | 2789.0 | 4 | 0.00 |
| null | -160.6 | 22.2 | 2834.3 | 5 | 0.00 |
| **Nigeria** | | | | | |
| mixture | - | - | 16178.3 | 1 | 1.00 |
| base | -138.0 | 18.0 | 16454.4 | 2 | 0.00 |
| null | -425.4 | 31.6 | 17029.0 | 3 | 0.00 |
| offset | -609.8 | 30.3 | 17397.9 | 4 | 0.00 |
| **Rwanda** | | | | | |
| base | - | - | 16.3 | 1 | 0.37 |
| null | -0.3 | 0.5 | 16.9 | 2 | 0.27 |
| mixture | -0.4 | 0.2 | 17.2 | 3 | 0.24 |
| mixture_offset | -1.6 | 1.0 | 19.5 | 4 | 0.07 |

*(Continued on Next Page...)*

Table S1: *(continued)*

| model | ELPD diff | SE | LOO-IC | rank | weight |
|---|---|---|---|---|---|
| offset | -2.2 | 1.8 | 20.7 | 5 | 0.04 |
| **Senegal** | | | | | |
| base | - | - | 183.9 | 1 | 0.43 |
| mixture | -0.3 | 0.5 | 184.4 | 2 | 0.33 |
| null | -0.6 | 1.9 | 185.0 | 3 | 0.24 |
| mixture_offset | -10.2 | 3.6 | 204.3 | 4 | 0.00 |
| offset | -26.3 | 6.5 | 236.6 | 5 | 0.00 |
| **Sierra Leone** | | | | | |
| mixture | - | - | 314.6 | 1 | 0.97 |
| mixture_offset | -3.6 | 6.1 | 321.8 | 2 | 0.03 |
| base | -5.2 | 4.5 | 325.0 | 3 | 0.01 |
| offset | -16.5 | 9.1 | 347.6 | 4 | 0.00 |
| null | -22.4 | 10.5 | 359.4 | 5 | 0.00 |
| **Somalia** | | | | | |
| mixture_offset | - | - | 1369.3 | 1 | 0.67 |
| mixture | -0.7 | 8.3 | 1370.8 | 2 | 0.32 |
| base | -5.2 | 8.8 | 1379.6 | 3 | 0.00 |
| offset | -14.2 | 10.5 | 1397.7 | 4 | 0.00 |
| null | -35.8 | 13.4 | 1440.8 | 5 | 0.00 |
| **South Africa** | | | | | |
| mixture_offset | - | - | 159.7 | 1 | 1.00 |
| base | -13.1 | 8.4 | 185.8 | 2 | 0.00 |
| mixture | -13.9 | 8.5 | 187.5 | 3 | 0.00 |
| null | -23.6 | 8.9 | 206.9 | 4 | 0.00 |
| offset | -24.0 | 10.0 | 207.7 | 5 | 0.00 |
| **South Sudan** | | | | | |
| mixture | - | - | 1102.6 | 1 | 1.00 |
| offset | -9.6 | 7.8 | 1121.8 | 2 | 0.00 |
| base | -25.9 | 7.4 | 1154.3 | 3 | 0.00 |
| null | -47.6 | 10.8 | 1197.8 | 4 | 0.00 |
| mixture_offset | -1555.7 | 297.2 | 4213.9 | 5 | 0.00 |
| **Sudan** | | | | | |
| null | - | - | 184.0 | 1 | 0.51 |
| mixture | -0.3 | 1.7 | 184.6 | 2 | 0.37 |
| base | -1.5 | 2.0 | 187.0 | 3 | 0.11 |
| mixture_offset | -41.3 | 4.4 | 266.6 | 4 | 0.00 |
| offset | -169.4 | 10.9 | 522.7 | 5 | 0.00 |
| **Tanzania** | | | | | |
| offset | - | - | 1763.5 | 1 | 1.00 |
| mixture_offset | -31.3 | 71.3 | 1826.0 | 2 | 0.00 |
| mixture | -71.5 | 21.7 | 1906.4 | 3 | 0.00 |
| base | -78.0 | 21.8 | 1919.4 | 4 | 0.00 |
| null | -97.5 | 27.5 | 1958.5 | 5 | 0.00 |
| **Tchad** | | | | | |
| mixture | - | - | 625.7 | 1 | 1.00 |
| base | -28.2 | 7.2 | 682.1 | 2 | 0.00 |
| mixture_offset | -41.4 | 11.2 | 708.5 | 3 | 0.00 |
| null | -78.8 | 13.4 | 783.3 | 4 | 0.00 |
| offset | -116.9 | 14.0 | 859.5 | 5 | 0.00 |
| **Togo** | | | | | |
| null | - | - | 410.7 | 1 | 0.93 |
| mixture | -3.0 | 1.4 | 416.6 | 2 | 0.05 |
| base | -4.3 | 1.3 | 419.3 | 3 | 0.01 |
| offset | -4.3 | 8.4 | 419.3 | 4 | 0.01 |
| mixture_offset | -178.3 | 77.8 | 767.3 | 5 | 0.00 |
| **Uganda** | | | | | |
| mixture | - | - | 2266.7 | 1 | 1.00 |
| base | -17.5 | 7.9 | 2301.7 | 2 | 0.00 |
| null | -24.3 | 11.8 | 2315.3 | 3 | 0.00 |
| offset | -68.8 | 20.5 | 2404.2 | 4 | 0.00 |
| mixture_offset | -1940.6 | 237.4 | 6147.9 | 5 | 0.00 |
| **Zambia** | | | | | |
| mixture_offset | - | - | 140.4 | 1 | 0.99 |
| base | -4.9 | 2.1 | 150.3 | 2 | 0.01 |
| mixture | -6.0 | 2.4 | 152.5 | 3 | 0.00 |
| null | -6.8 | 3.0 | 154.0 | 4 | 0.00 |
| offset | -7.5 | 3.4 | 155.4 | 5 | 0.00 |
| **Zimbabwe** | | | | | |
| mixture | - | - | 704.6 | 1 | 0.94 |
| base | -2.7 | 1.6 | 710.0 | 2 | 0.06 |
| null | -14.4 | 6.2 | 733.5 | 3 | 0.00 |
| mixture_offset | -28.9 | 19.5 | 762.4 | 4 | 0.00 |
| offset | -33.5 | 10.9 | 771.6 | 5 | 0.00 |

#### S4 Posterior predictive checks

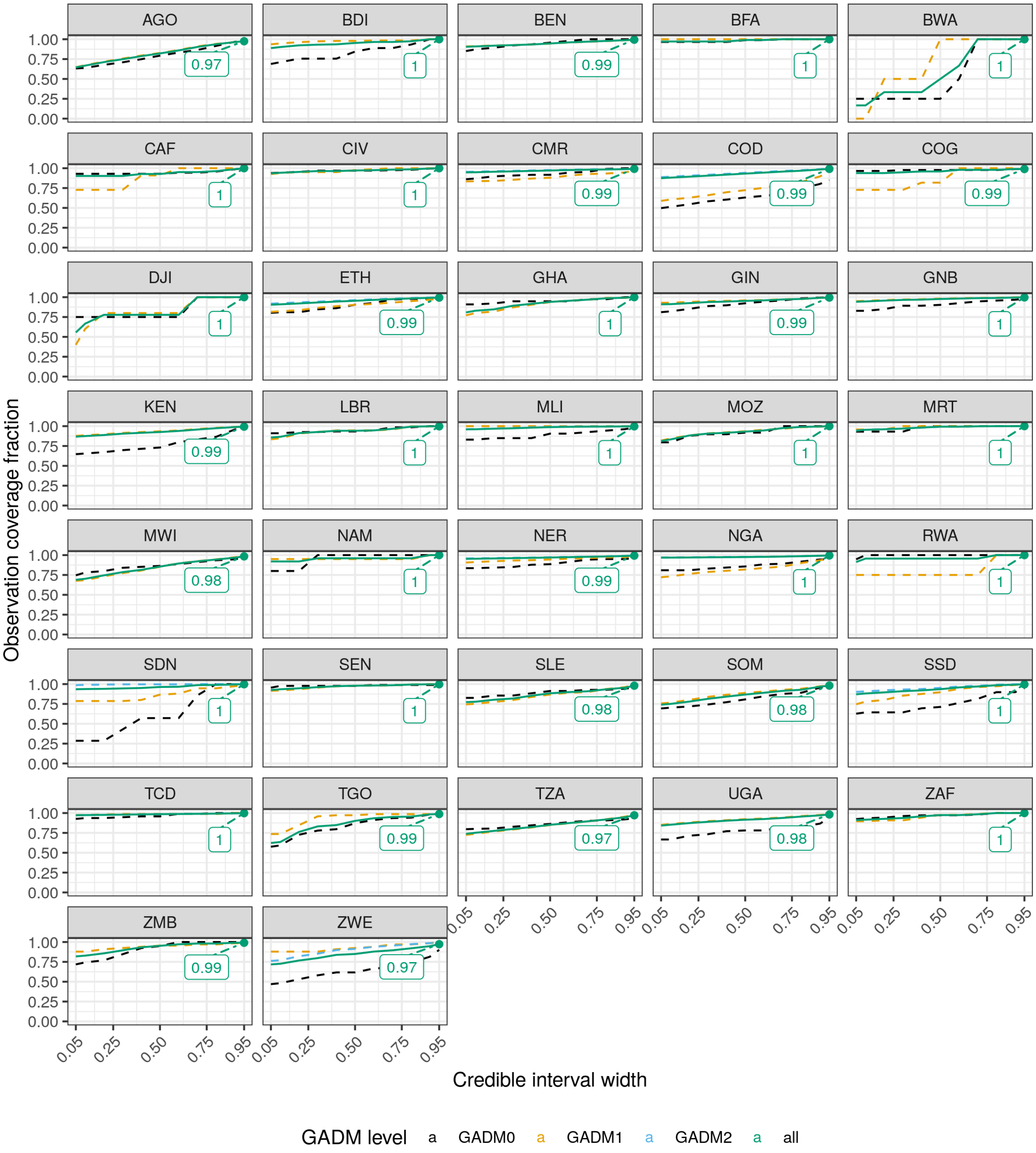


Figure S2: Model evaluation. For each country the performance of the selected model is given as the fraction of observations covered by posterior predictions as a function of the credible interval width. The coverage fraction is given for all observations as well as partitioned by administartive level.

### S5 Additiona results

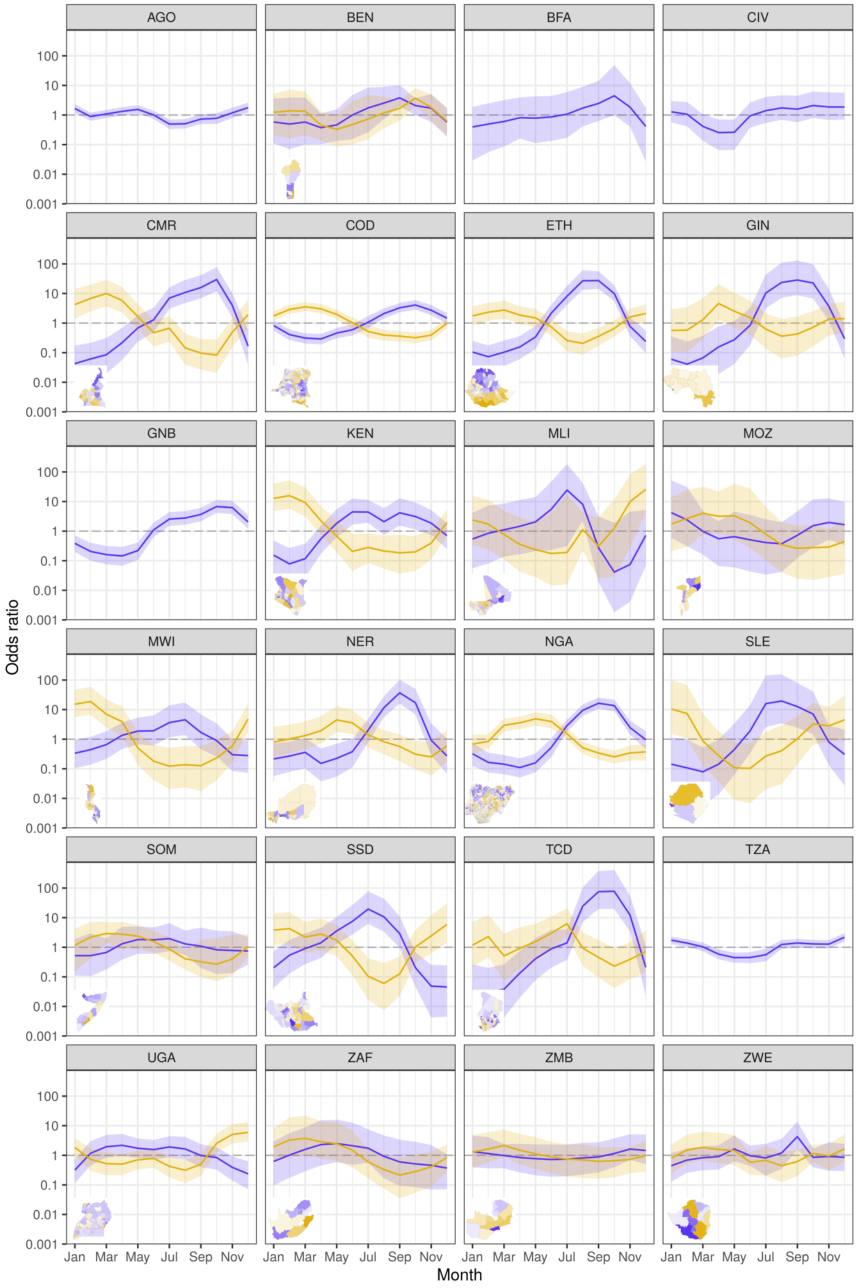


Figure S3: Country-level cholera sesaonality time series. Excess cholera seasonality is shown in for the final selection of models in terms of the mean (line) and 95% CrI odds ratios.

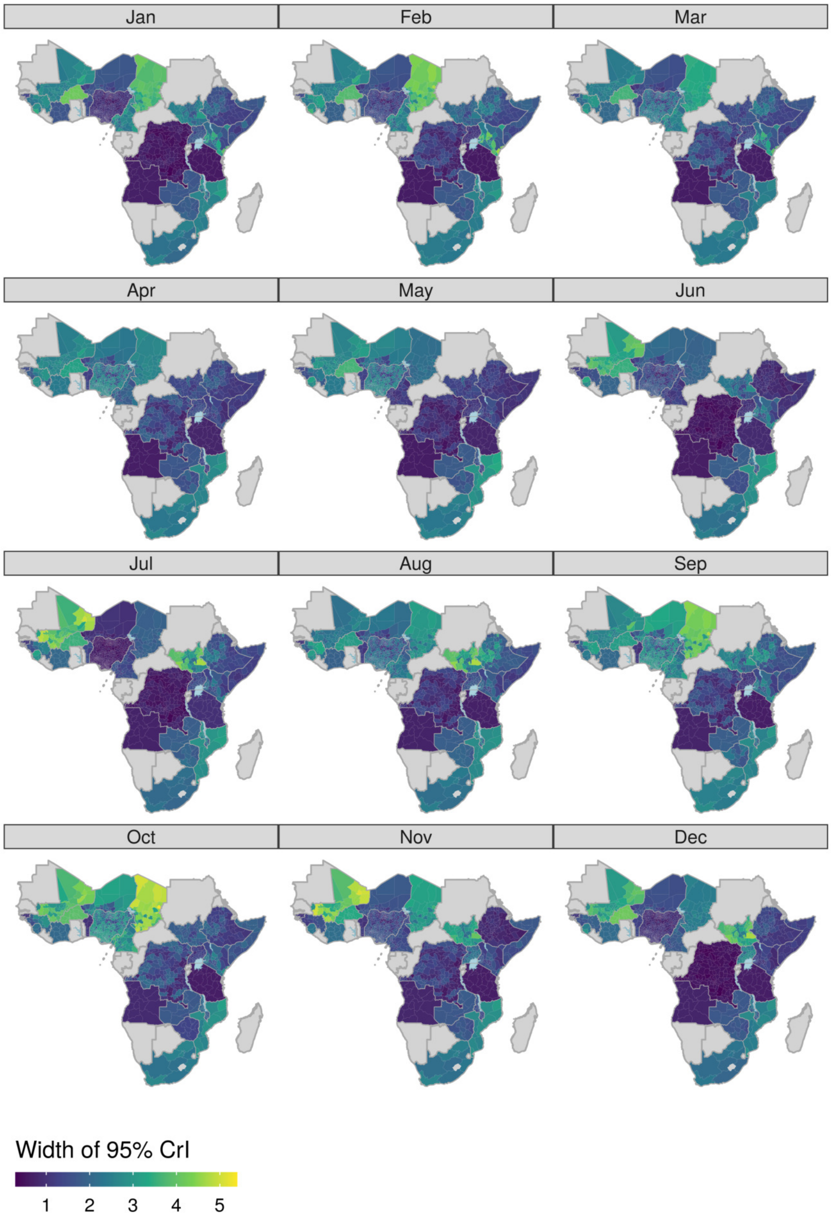


Figure S4: Uncertainty in seasonality coefficients in terms of the width of the 95 % CrI on the logit scale.

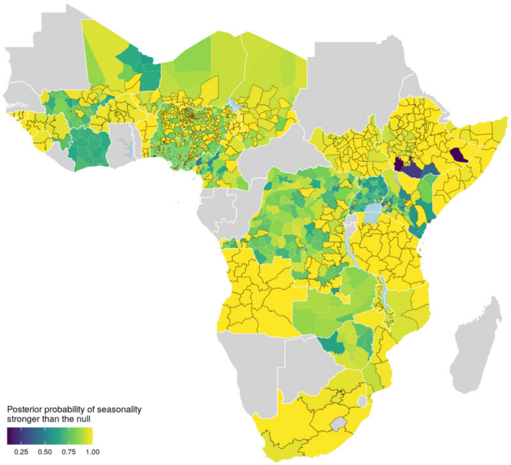


Figure S5: Uncertainty in seasonality index in terms of the posterior probability that the index is larger than the null hypothesis of equal occurrence probability during the year. Adminstrative units for which the posterior probability is above 0.9 are outlined in black.

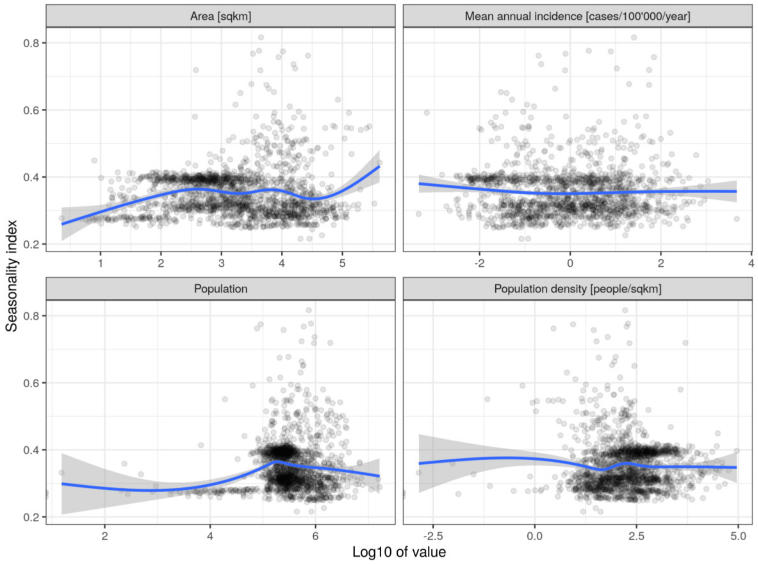


Figure S6: Seasonality strength and administrative unit characteristics. Each point corresponds to one administrative unit, lines correspond to GAM estimates of the relationship betwen the characteristics and the seasonality indeces.

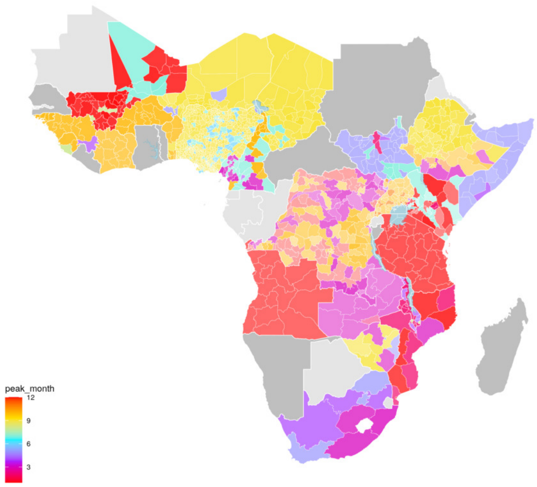


Figure S7: Seasonality peak map. Map of the month with largest estimated excess cholera occurrence odds ratio in countries for which a model with seasonality was selected.

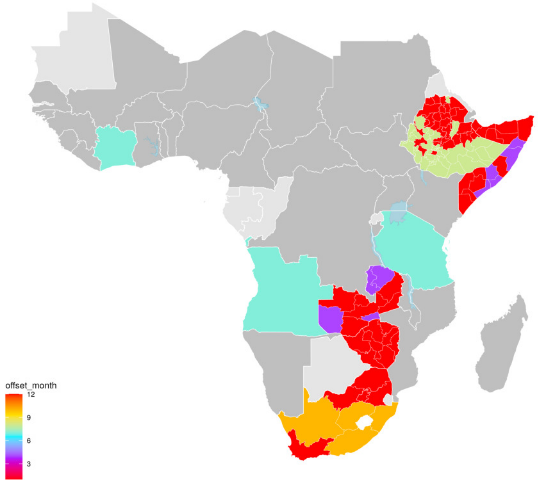


Figure S8: Offset map. Map of the month determining the start of the 'cholera year' in coutries for which a model with offset was selected. Countries with two colors correspond to models with both mixture and offset.

#### S6 Seasoanlity grouping

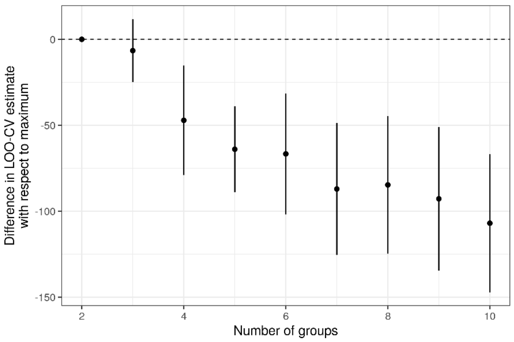


Figure S9: Model comparison of number of groupings. Model comparison was done in terms of the estimated log-predictive density (elpd). Pairwise differences in elpd were computed for all model combinations, and results are shown in terms of the mean (points) and 2 standard errors (lines). The model with highest elpd was the model with 2 groups.

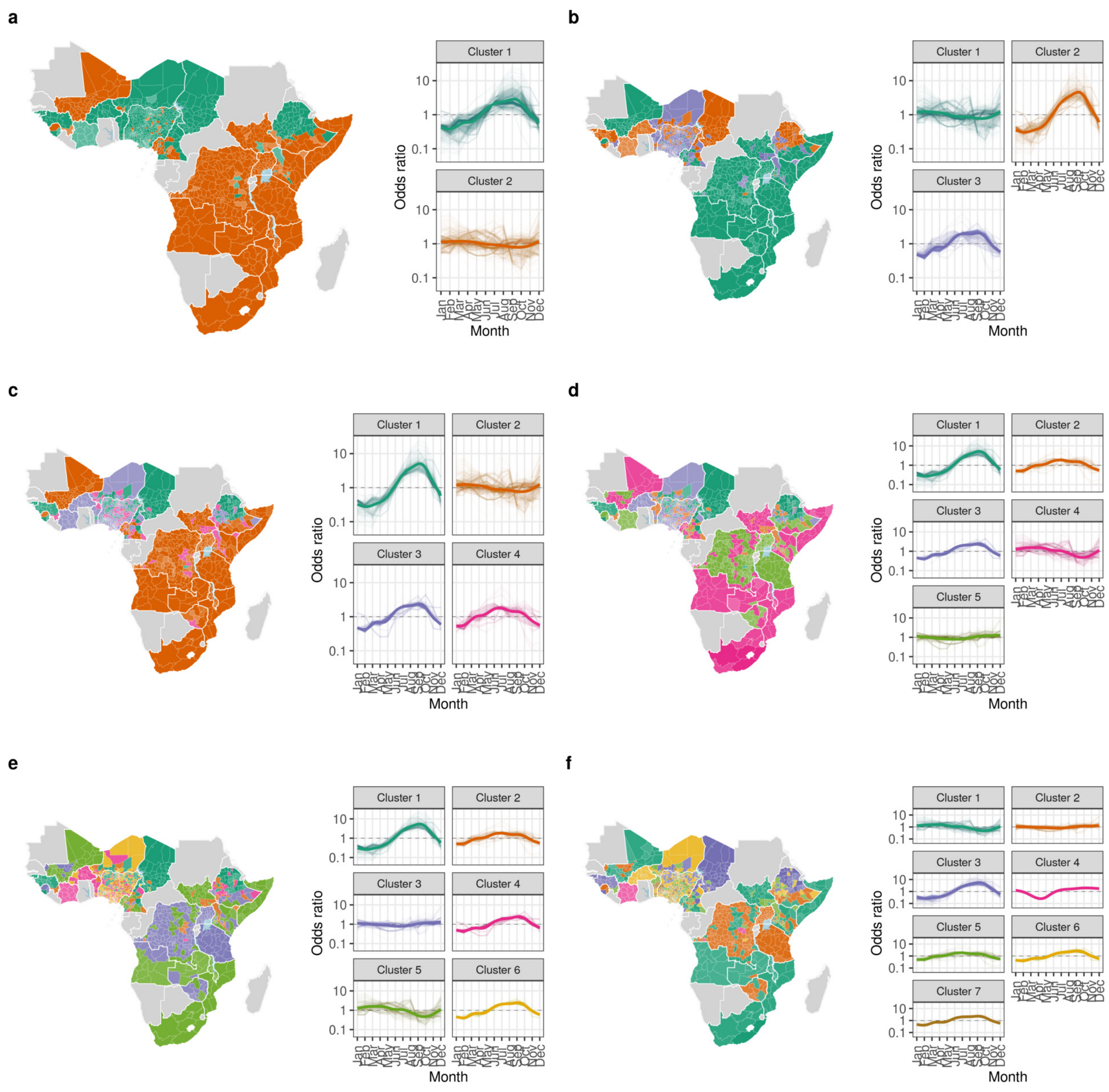


Figure S10: Clustering results for two to seven clusters.

#### S7 Correlation with hydro-climatic variables

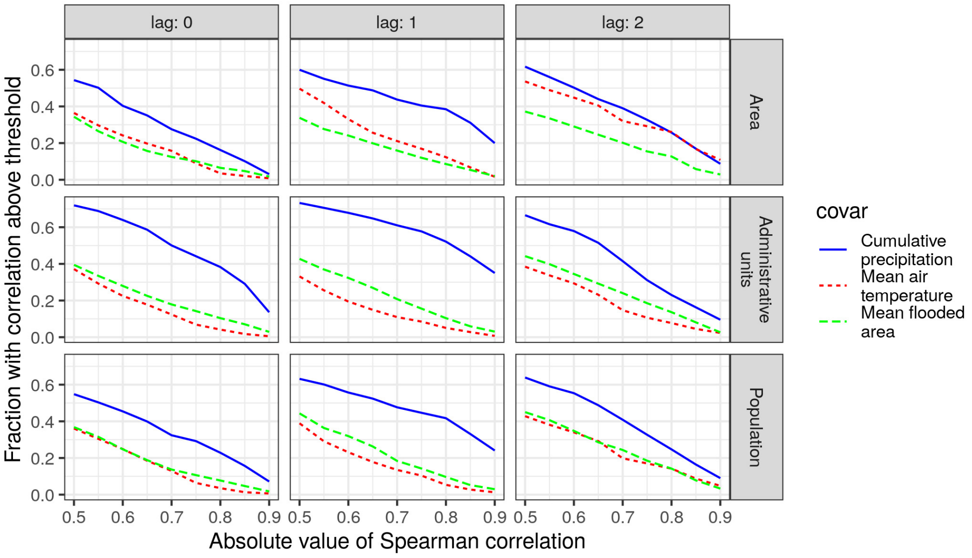


Figure S11: Importance of correlations with hydro-climatic variables. Importance is given in terms of the fraction of area, administrative units, and population with the absolute value of Spearman correlation above thresholds between 0.5 and 0.9.

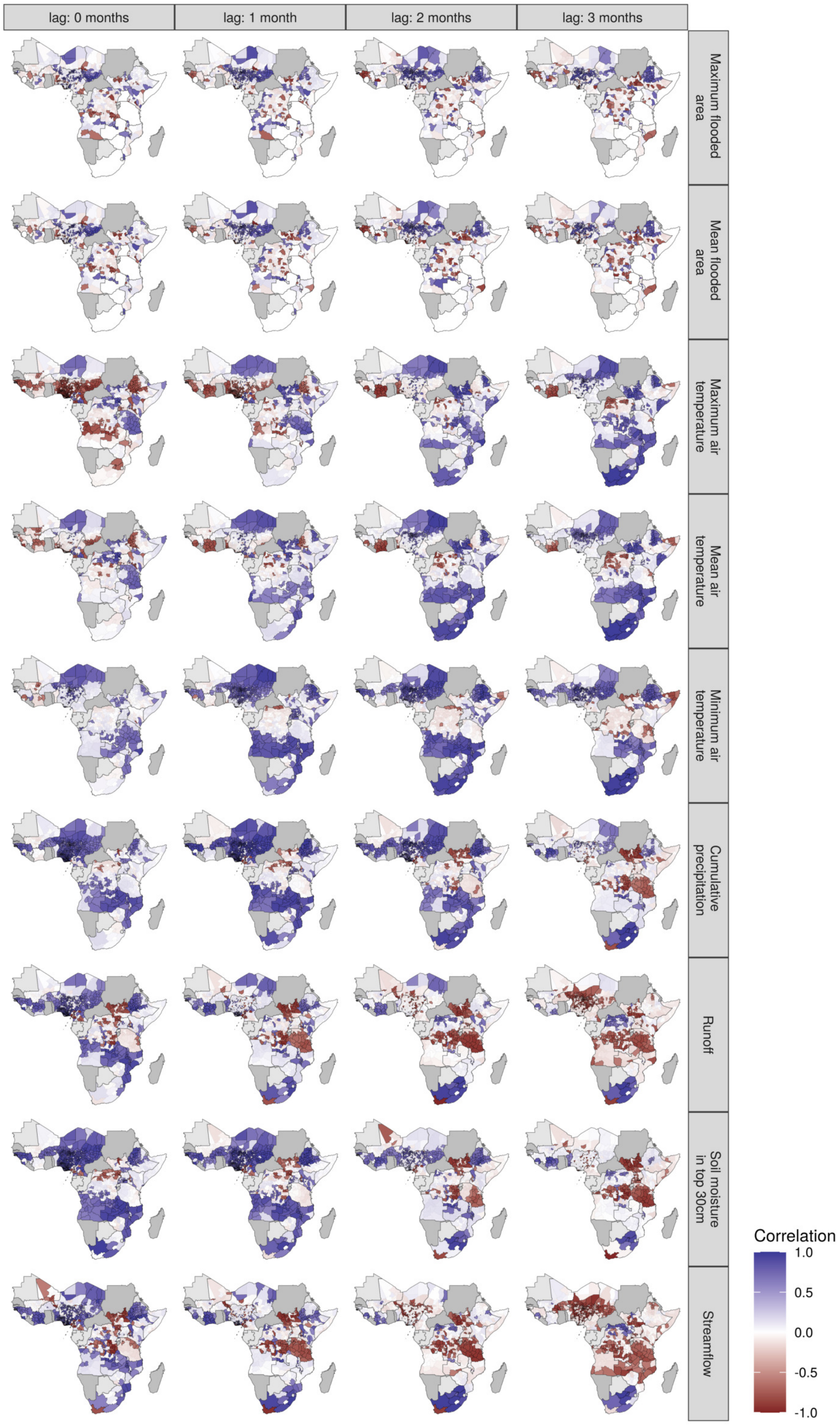


Figure S12: Correlation between excess cholera seasonality and hydro-climatic variables. Correlation is experssed in terms of Spearman's coefficient, with areas with significant correlations outlined in balck.

#### S8 Alternative cholera occurrence definitions

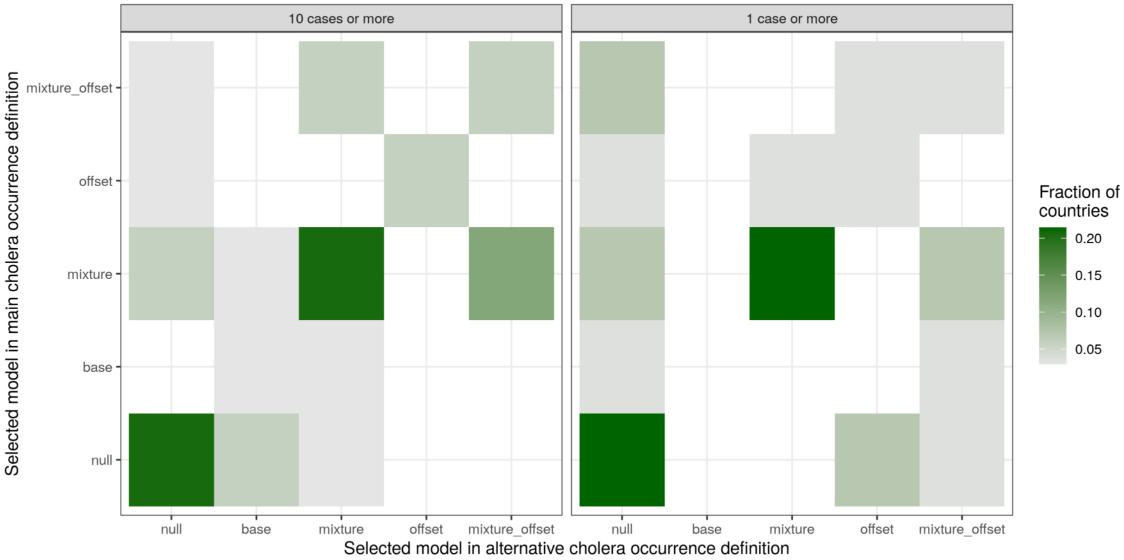


Figure S13: Comparison of model selection for alternative definitions of cholera occurrence
